# Extracorporeal Fixation and Re-implantation for High Condylar Split Fractures: The Motamed Technique

**DOI:** 10.64898/2026.06.02.26354404

**Authors:** Ahmed Abdul-Motamed Amer

## Abstract

**Background:** Management of high sagittal split fractures of the mandibular condyle remains a formidable surgical challenge due to limited visualization, technical difficulties in direct in-situ fixation, and the high risk of secondary avascular necrosis or temporomandibular joint (TMJ) ankylosis.

**Objectives:** To evaluate the clinical outcomes and technical efficacy of the Motamed Technique, a standardized protocol involving extracorporeal rigid internal fixation followed by anatomical re-implantation for complex high condylar split fractures.

**Methods:** **A** retrospective evaluation was conducted on a clinical series of 11 **consecutive patients (9 males, 2 females)** presenting with severe, displaced high sagittal split condylar fractures secondary to high-velocity trauma. In all cases, the fragmented condylar segments were completely retrieved, stabilized ex vivo on a back-table using a titanium X-shaped 3D mini-plate system (1.5 mm), and meticulously re-implanted into the glenoid fossa. Total cold ischemia time was strictly maintained between 10 and 20 minutes. **The postoperative longitudinal follow-up period ranged from 6 to 11 months (mean duration: 8.6 months)**. Comprehensive post-operative tracking included clinical parameter checking, 3D Computed Tomography (3D-CT), and high-resolution dynamic Magnetic Resonance Imaging (MRI) to analyze bony union, vertical ramus height restoration, and articular disc kinetics.

**Results:** All 11 patients achieved predictable and stable functional outcomes. At the definitive milestones, the mean maximum mouth opening **(MMO)** was **37.3 mm (range, 33** – **45 mm)**, demonstrating excellent vertical clearance and stable lateral/protrusive excursions. Pre-traumatic stable centric occlusion was perfectly restored and maintained in **100% of cases (n=11)**, with zero incidence of postoperative open bite or crossbite. Facial nerve motor function was entirely preserved across the cohort **(100% House-Brackmann Grade** I at final follow-up). Longitudinal 3D-CT scans confirmed complete osseous union and anatomical alignment in all cases by the 4th postoperative month, with no radiographic evidence of condylar head resorption or hardware failure. Follow-up MRI findings demonstrated the preservation of TMJ dynamics, functional articular disc movement (with stable reduction in 3 cases), and a total absence of avascular necrosis or intra-articular effusion. No cases of TMJ ankylosis were reported.

**Conclusion:** The Motamed Technique provides a reliable, reproducible, and biologically sound approach for managing intricate high condylar split fractures. By utilizing systematic extracorporeal mini-fixation, this protocol effectively overcomes intraoperative spatial limitations while ensuring excellent long-term anatomical stability, stable occlusion, and functional joint mobility without compromising facial nerve integrity.

## 1. Introduction

Mandibular condylar fractures represent a substantial proportion of maxillofacial traumas, accounting for approximately 25% to 35% of all mandibular injuries [1]. Among these, high sagittal split fractures of the condylar head present an exceptional therapeutic dilemma for oral and maxillofacial surgeons. The intimate anatomical relationship with the temporomandibular joint (TMJ) capsule, the proximity of the delicate branches of the facial nerve, and the severely restrictive intraoperative visualization often preclude precise anatomical reduction through conventional open reduction and internal fixation (ORIF) in-situ [2]. The surgeon is frequently forced to choose between inadequate fixation due to limited physical access or risking excessive soft-tissue retraction, which heavily jeopardizes the regional neurovascular structures.

Historically, closed functional management with maxillomandibular fixation (MMF) was advocated to bypass these surgical morbidities. However, contemporary long-term cohort data have unequivocally demonstrated that conservative approaches for highly displaced fractures frequently culminate in severe secondary sequelae, including diminished ramal height, persistent malocclusion, chronic TMJ pain, and internal joint derangement [3]. Conversely, attempting direct in-situ fixation within the deep, narrow confines of the glenoid fossa often leads to poorly oriented hardware placement, failure to achieve rigid stability, and a high incidence of secondary displacement.

To resolve these intraoperative structural limitations, the concept of extracorporeal fixation and re-implantation–wherein the proximal fragmented segments are deliberately retrieved, reconstructed ex-vivo, and subsequently replanted–has emerged as a logical alternative [4]. By converting a deeply buried, obscured fracture into an open, accessible ex-vivo procedure, the surgeon can achieve absolute anatomical precision.

Nevertheless, this aggressive approach introduces a distinct biological paradox. Complete removal of the condylar segments results in total stripping of the periosteum and the lateral pterygoid muscle attachment, completely disrupting the primary nutrient vascular supply derived from the maxillary artery and its branches. A comprehensive systematic review published recently by Al-Ghamdi et al. (2025) highlighted that while extracorporeal fixation provides an unparalleled benefit for achieving precise marginal alignment in inaccessible fractures, it traditionally harbors a critical baseline concern regarding secondary avascular necrosis, progressive condylar resorption, or eventual fibrous/bony TMJ ankylosis [5]. The success of this procedure hinges entirely on a delicate race between creeping substitution (bony remodeling) and osteoclastic bone resorption.

Furthermore, contemporary consensus frameworks, such as the 13-year institutional review by Raimondo et al. (2026), emphasize that establishing structural osseous union alone is insufficient; advanced high-resolution imaging (3D-CT and MRI) is imperative to prove that structural re-implantation translates into true physiological and dynamic joint recovery, characterized by functional articular disc mobility and the absence of chronic intra-articular effusion [6].

while various stabilization methods have been historically described, a critical clinical gap persists regarding a standardized protocol tailored specifically for highly displaced, comminuted high sagittal split condylar fractures that consistently guarantees long-term dynamic joint health without risking avascular complications. Traditional extracorporeal approaches often suffer from prolonged cold ischemia times or rely on micro-fixation systems that lack the multi-directional biomechanical strength required to withstand early functional loading, thereby accelerating micro-motion and bone resorption.

To bridge this specific clinical and technical dilemma, this paper introduces the **“Motamed Technique”** – a refined, highly systematic surgical protocol combining a modified deep retromandibular transparotid approach with rapid ex-vivo micro-reduction, rigid titanium X-plate fixation, and immediate structural re-implantation. Rather than presenting an isolated technical note or an un-reproducible methodology, this clinical article documents the therapeutic significance of this protocol through a consecutive series of 11 **patients** tracked via a comprehensive **longitudinal follow-up period spanning from** 6 to 11 **months**. Utilizing rigorous postoperative 3D-CT and high-resolution dynamic MRI, this study aims to demonstrate that the Motamed Technique predictably delivers absolute anatomical stability, fully stable occlusion, complete preservation of facial nerve integrity, and long-term physiological TMJ dynamics with functional disc movement.

### Study Design and Ethical Approval

This retrospective clinical investigation was formally reviewed and approved by the Institutional Review Board (IRB) and the Research Ethics Committee of the Craniomaxillofacial Surgery Department at Sherbin Central Hospital, Dakahlia, Egypt. Formal ethical approval and administrative clearance were officially granted under the specific institutional registration number: SCH-2025-OMFS-094.

### Patient Cohort and Selection Criteria

The expanded study cohort comprised *9* male and 2 female patients, presenting with a mean age of 25.8 years (range, 19–35 years). All fractures were sustained secondary to high-velocity road traffic accidents (RTAs) or severe falls.

- **Inclusion Criteria:** Skeletally mature adult patients (age 18 years); presentation with severely displaced or dislocated isolated high sagittal split condylar head fractures, or complex cases accompanied by secondary maxillofacial fractures (symphyseal, parasymphyseal, body, angle, or midface fractures); and an unremarkable medical history.
- **Exclusion Criteria:** Patients aged >60years; systemic metabolic bone diseases (e.g., osteoporosis); uncontrolled chronic medical conditions; or failure to comply with the longitudinal follow-up regimen.

### Anatomical Fracture Distribution

The anatomical distribution of the fractures demonstrated a high level of trauma complexity:

- **Bilateral Condylar Head Fractures (n=3):** Fractures were accompanied by complex mandibular symphyseal/parasymphyseal fractures or extensive panfacial/nasal bone configurations.
- Unilateral Condylar Head Fractures (n=8): Fractures were associated with secondary maxillofacial injuries, including associated configurations of mandibular body/angle fractures, zygomaticomaxillary complex (ZMC) disruptions, and Le Fort midface fracture patterns.

### Preoperative Protocol and Prophylactic Regimen

One hour prior to the surgical intervention, all patients received a strict prophylactic intravenous regimen to minimize postoperative infection and edema. This consisted of 2 grams of Cefazolin and 4 mg of Dexamethasone diluted in a 500 mL normal saline infusion (0.9% NaCl), administered intravenously. A mandatory skin hypersensitivity test was performed for all patients preoperatively to rule out any allergic reactions to the antibiotic.

### Postoperative Evaluation Framework

A standardized, longitudinal clinical and radiographic follow-up regimen was uniformly implemented for the entire cohort. Preoperative assessment utilized high-resolution 3D computed tomography (3D-CT) to map the fracture lines and quantify fragment displacement. Postoperatively, longitudinal evaluations were conducted at terminal milestones tailored to each patient’s timeline. **The definitive postoperative follow-up period ranged from 6 to 11 months, yielding a mean follow-up duration of 8.6 months**. Specifically, 3 patients completed an 11-month follow-up, 4 patients completed a 9-month follow-up, and 4 patients reached the 6-month milestone.

Clinical tracking verified stable centric occlusion, maximum mouth opening (MMO), and lateral/protrusive jaw excursions. Facial nerve motor function was systematically graded using the House-Brackmann classification system. Radiographic evaluations at the terminal milestones utilized high-resolution 3D-CT scans to verify solid osseous union, and high-resolution, non-contrast dynamic Magnetic Resonance Imaging (MRI) to investigate temporomandibular joint (TMJ) dynamics, articular disc translation, and to definitively rule out secondary avascular necrosis or chronic joint effusion.

### Statistical Analysis

Categorical and continuous variables were analyzed using descriptive statistical methods. Quantitative metrics, including patient chronological age, duration of follow-up, and postoperative maximum mouth opening (MMO), were computed as means, standard deviations (SD), and ranges. Qualitative parameters–such as anatomical fracture distribution, gender, occlusal stability, and neurological status (House–Brackmann grades)–were reported as frequencies and percentages. All statistical computations were executed using SPSS Software (Version 26.0; IBM Corp, Armonk, NY, USA).

### Surgical Technique: The Motamed Technique

#### Surgical Exposure and Facial Nerve Preservation

Under standard general anesthesia via nasotracheal intubation, open access to the fractured condylar region was achieved. A standard retromandibular transparotid approach was performed on the affected side(s). The incision was placed approximately 0.5 to 1 cm posterior to the posterior border of the mandibular ramus. Dissection proceeded carefully through the subcutaneous tissues and the capsule of the parotid gland.

Anterograde dissection was meticulously carried out through the deep portion of the parotid gland (deep transparotid layer). The main trunk and the relevant branches of the facial nerve–specifically the marginal mandibular, buccal, and temporal branches–were identified, mapped, and fully preserved using blunt, atraumatic dissection. In cases where high-velocity trauma had already resulted in a traumatic open wound in the regional tissue, this existing entry point was strategically utilized and incorporated into the surgical exposure to minimize further tissue trauma. No external counter-incisions were performed, ensuring that the entire intra-articular and ramal stabilization remained controlled internally. Following safe nerve retraction, the pterygomasseteric sling was incised, and the masseter muscle was elevated to achieve direct, comprehensive exposure of the fractured condylar unit and the joint capsule. Depending on the clinical presentation, this precise approach was applied systematically to either unilateral or bilateral high sagittal split fractures.

#### Atraumatic Retrieval and Ex-Vivo Fixation

The severely displaced and telescoped high sagittal split fragments were gently isolated from the surrounding soft tissues and retrieved from the glenoid fossa with fine instruments. Special care was taken to avoid any forceful manual manipulation within the deep space to prevent neurovascular injury. The retrieved bone fragments were immediately transferred to a sterile back-table workstation for ex-vivo bench stabilization. To preserve osteocyte and chondrocyte viability, the segments were completely immersed in a chilled isotonic saline solution enriched with broad-spectrum antibiotics, maintaining a protected cold ischemic environment.

Under direct, unhindered 360-degree visualization and optimal lighting on the back-table, the split sagittal segments were anatomically approximated using micro-reduction forceps to perfectly restore the exact pre-traumatic contours and morphology of the condylar head. Rigid internal fixation of the split fragments was achieved ex-vivo utilizing a low-profile titanium X-shaped 3D mini-plate system (1.5 mm). The X-plate was secured across the cross-fracture lines using 7 mm monocortical screws, providing superior biomechanical multi-directional stability and anti-torsional resistance without penetrating the functional intra-articular surface. Due to the efficiency of the standardized back-table protocol, the total cold ischemia time strictly ranged between 10 to 20 minutes across all cases.

#### Occlusal Stabilization, Re-implantation, and Ramal Fixation

Prior to re-implanting and securing the reconstructed condyle to the mandibular ramus, the establishment of intermaxillary fixation (IMF) was performed as a critical, mandatory initial step. The teeth were placed and rigidly locked into their precise, pre-traumatic centric occlusion utilizing IMF screws or intermaxillary wires. This maneuver was executed to secure baseline skeletal stability and to definitively lock the correct vertical ramus height symmetrically before anchoring the joint structure.

Once the maxillomandibular relationship was rigidly locked, the extracorporeally stabilized condylar unit was transferred back into the surgical field and re-implanted precisely into its native anatomical position within the glenoid fossa, ensuring an optimal disc-graft relationship. The reconstructed condylar base was perfectly aligned with the distal segment of the mandibular ramus and rigidly secured utilizing a combination of positional and adaptation continuous posterior miniplates to effectively withstand functional muscular vectors.

#### Closure and Postoperative Rehabilitation

The temporary MMF wires were released intraoperatively to manually confirm smooth, unhindered mandibular translation and rotation without joint catching or mechanical lock. The TMJ capsule was meticulously re-approximated and sutured. Layered closure of the fascia, subcutaneous tissues, and skin was performed. Postoperatively, non-rigid MMF elastics were maintained for two weeks (14 days) to guide and secure the occlusion during early healing. This was followed by the immediate initiation of a structured physical therapy and jaw-opening exercise protocol (4 to 6 times daily) to prevent intra-capsular adhesion formation and promote functional joint remodeling.

## 3. Results

**Figure 1:**
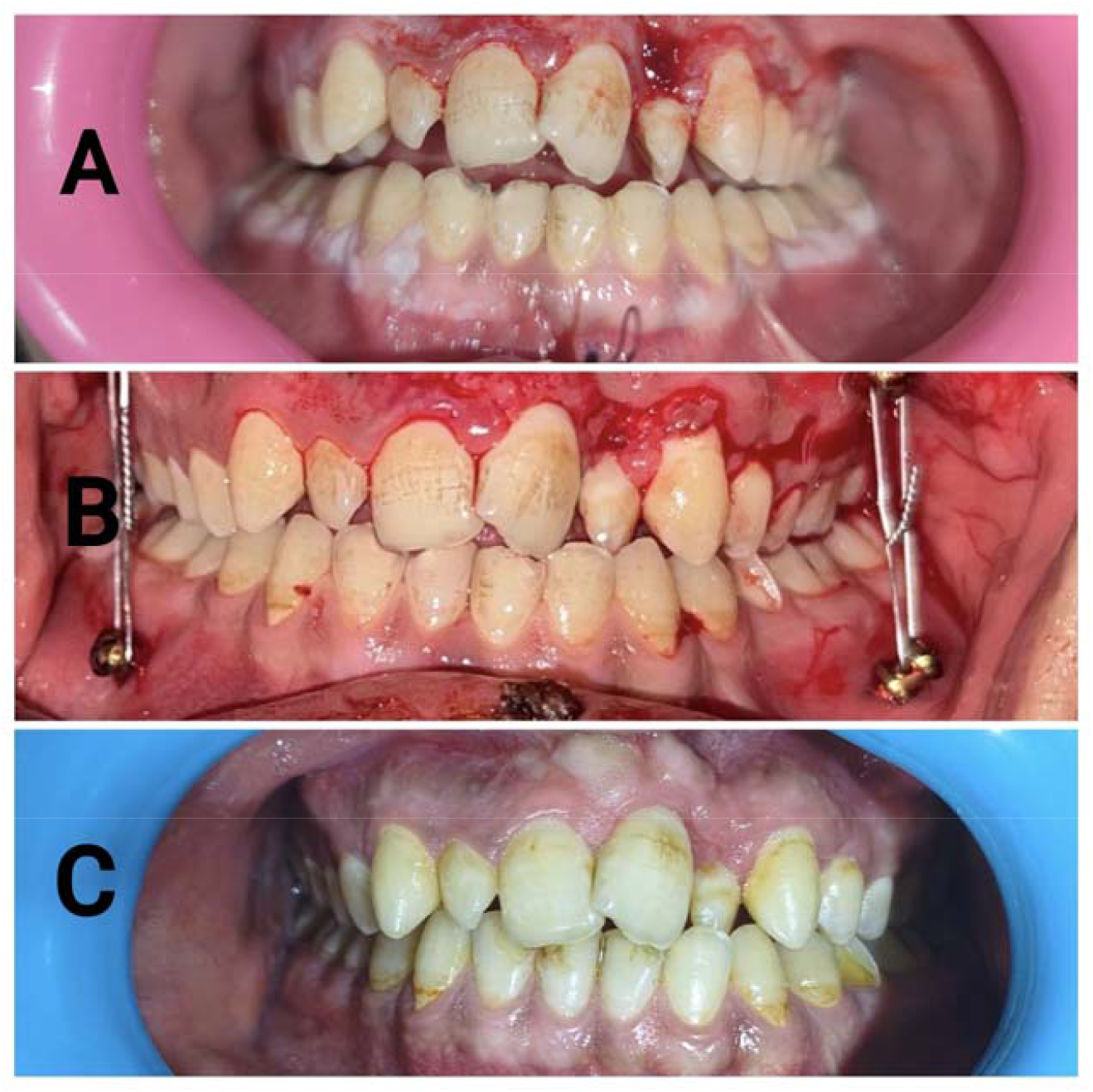
Serial clinical photographs demonstrating the management and occlusal rehabilitation of a high split condylar fracture. (A) Preoperative clinical view showing post-traumatic malocclusion, anterior open bite, and deviation resulting from the condylar displacement. (B) Intraoperative view showing the establishment of temporary maxillomandibular fixation (MMF) utilizing intermaxillary fixation (IMF) screws to restore proper occlusal relationships and guide anatomical condylar alignment. (C) At the definitive post postoperative follow-up (range, 6–11 months) demonstrating excellent long-term occlusal stability, complete resolution of the malocclusion, and successful functional rehabilitation of the mandibular movements.

**Figure 2:**
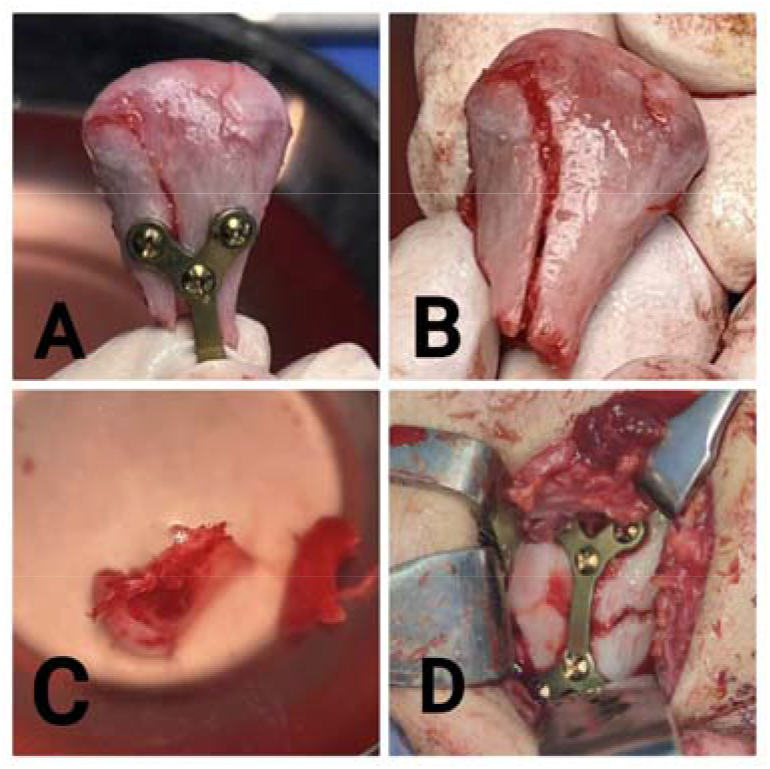
Intraoperative photographs illustrating the extracorporeal reduction, rigid internal fixation, and replantation of the high split condylar segment. (A) Extracorporeal anatomical reduction and rigid osteosynthesis of the fractured condylar fragments utilizing a titanium X-plate and mini-screws. (B) The retrieved condylar head demonstrating a vertical/sagittal split fracture before fixation. (C) Debridement and preparation of the articular fossa and joint space. (D) Successful replantation and anatomical positioning of the reconstructed condylar unit back into its native site, securing stable orientation and internal fixation.

**Figure 3:**
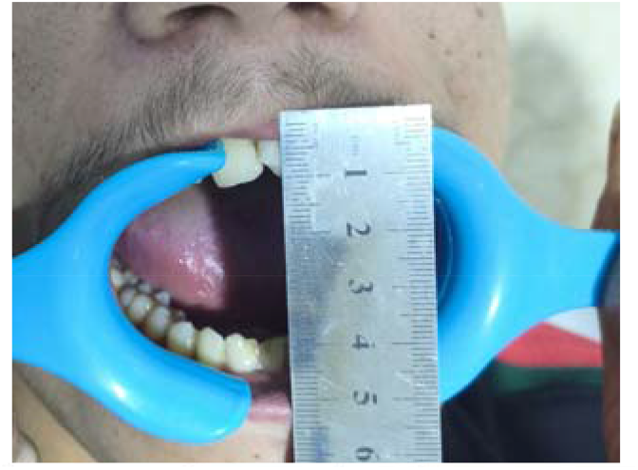
At the definitive postoperative follow-up (range, 6–11 months) Clinical photograph demonstrating an excellent maximum interincisal opening (MIO) of approximately 45 mm. This reflects complete functional rehabilitation, stable mandibular mobility, and no signs of temporomandibular joint (TMJ) ankylosis or restriction following the extracorporeal fixation of the high split condylar fracture.

**Figure 4:**
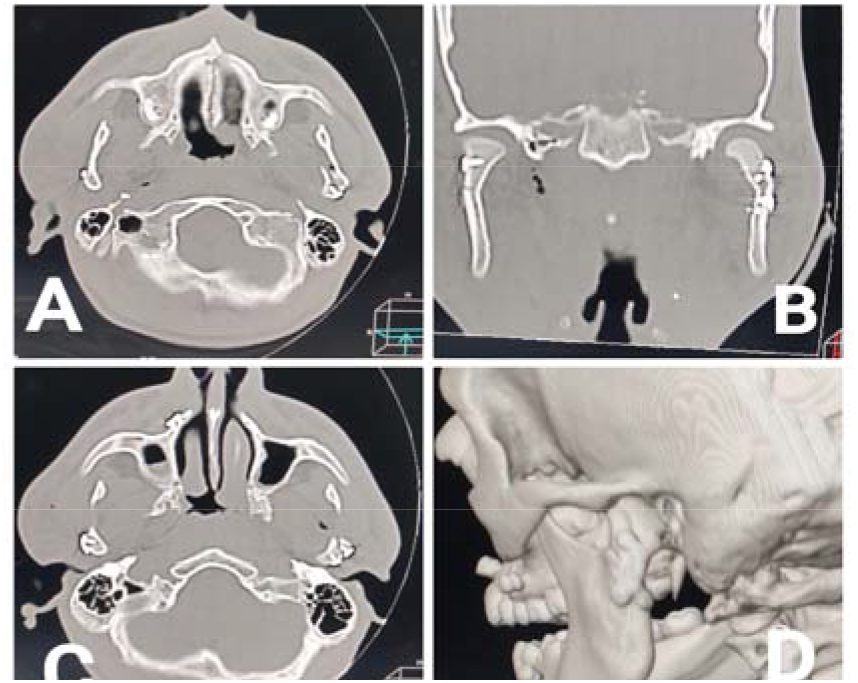
Immediate postoperative computed tomography (CT) scans confirming anatomical reconstruction. (A, C) Axial CT views showing correct alignment and stable position of the replanted condylar segments within the glenoid fossa. (B) Coronal CT view demonstrating precise vertical height restoration of the mandible and rigid internal fixation of the high split condylar fracture achieved by the titanium X-plate and micro-screws. (D) Three-dimensional (3D) CT reconstruction providing definitive evidence of excellent anatomical contouring, continuity, and structural integrity of the reconstructed condylar unit.

**Figure 5:**
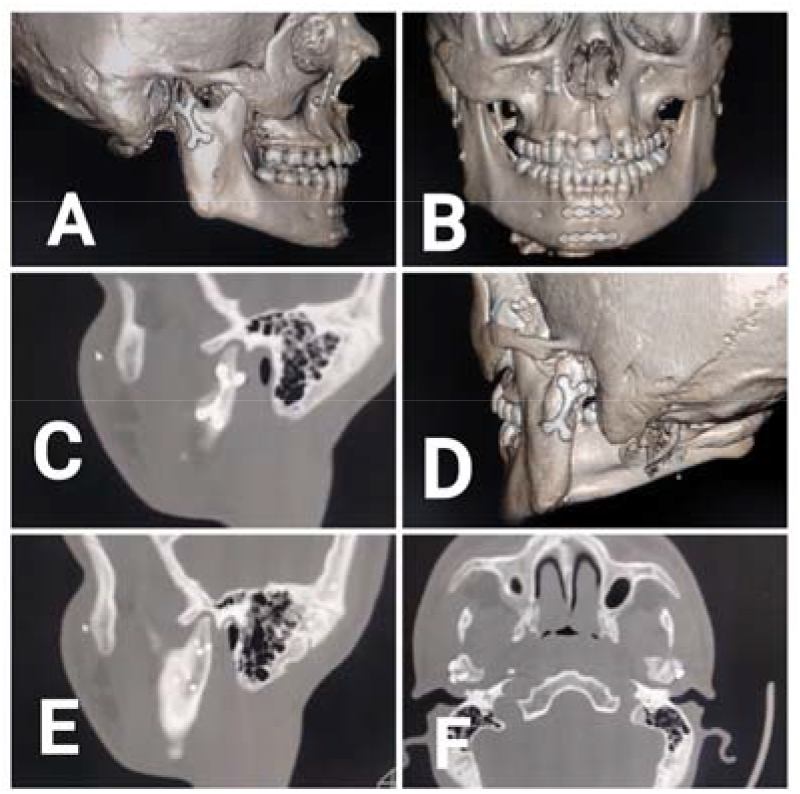
At the definitive postoperative follow-up (range, 6–11 months) computed tomography (CT) scans evaluating bone remodeling and fixation stability. (A, D) Three-dimensional (3D) CT reconstructions demonstrating excellent bony consolidation and complete remodeling of the reconstructed condylar unit, with stable positioning of the X-plate and screws. (B) Frontal 3D CT view showing optimal facial symmetry and maintenance of the corrected mandibular vertical height. (C, E) Sagittal CT views confirming solid osseous union across the fracture line and anatomical integrity of the condylar head within the articular eminence. (F) Axial CT view illustrating stable axial alignment of both condyles without any signs of osteolysis or hardware displacement.

**Figure 6:**
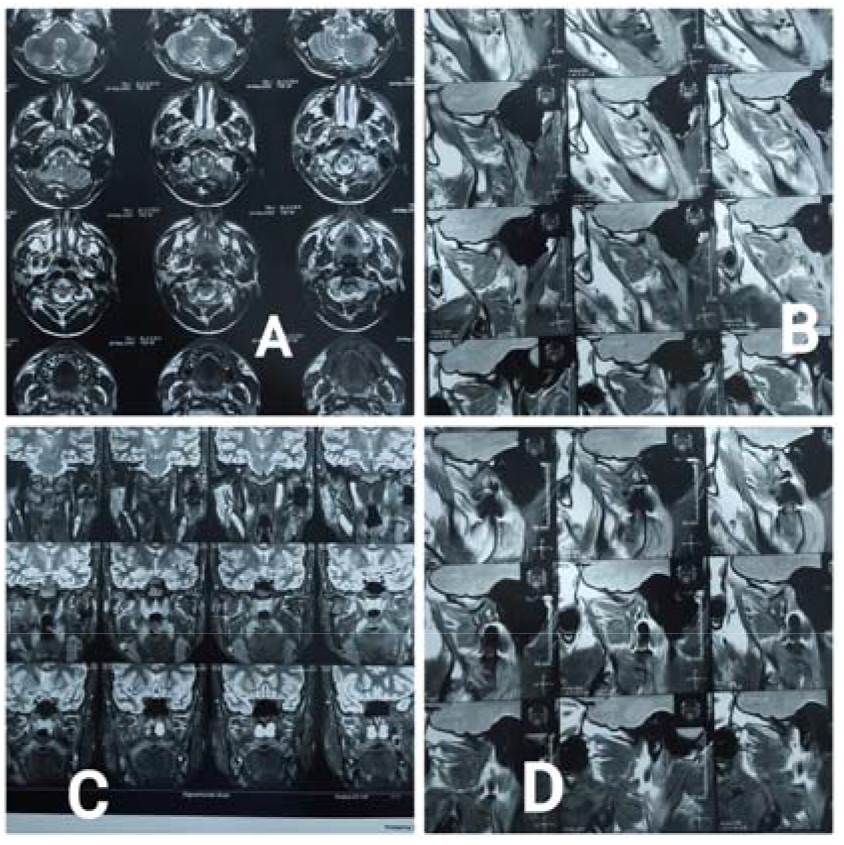
At the definitive postoperative follow-up (range, 6–11 months) magnetic resonance imaging (MRI) of the temporomandibular joints (TMJs) for comprehensive soft tissue and disc-status evaluation. (A, C) Axial and coronal T1 & T2-weighted MRI views demonstrating complete osseous consolidation of the replanted condyle, excellent periarticular tissue healing, and no detectable synovial effusion. (B, D) Sagittal MRI views in closed-mouth and open-mouth positions revealing a mild bilateral internal derangement characterized by anterior disc displacement with reduction (ADDWR). Concomitant normalization of the anatomic relationship between the mandibular condyle and the infratemporal bony eminence is definitively demonstrated during maximum mouth opening, confirming stable functional remodeling and restoration of smooth translational joint mobility.

### Patient Demographics and Fracture Distribution

A total of 11 consecutive patients (9 males and 2 females) with a mean age of 25.8 years (range, 19–35 years) were retrospectively evaluated. All cases presented with severely displaced or dislocated high sagittal split fractures of the mandibular condylar head sustained secondary to high-velocity road traffic accidents (RTAs) (n=9) or severe falls (n=2).

The anatomical distribution and trauma patterns across the cohort demonstrated a high level of maxillofacial complexity:

- **Bilateral Condylar Head Fractures (n=3):** One case presented with a concomitant mandibular symphyseal fracture, the second case with a mandibular parasymphyseal fracture combined with nasal bone fractures (panfacial pattern), and the third case involved bilateral condylar head fractures associated with a comminuted mandibular body fracture.
- **Unilateral Condylar Head Fractures (n=8)**: All eight cases were associated with secondary maxillofacial injuries, including associated combinations of mandibular fractures (body/angle) and complex midfacial fracture configurations (zygomaticomaxillary complex [ZMC] and Le Fort patterns).

#### Clinical and Functional Outcomes

At the definitive postoperative milestone assessments, remarkable functional restoration of the masticatory and stomatognathic system was achieved across the entire expanded cohort:

- **Maximal Mouth Opening (MMO):** The mean postoperative MMO achieved for the cohort was 37.3 mm (ranging precisely from 33 to 45 mm), representing an excellent clinical restoration of vertical mandibular clearance and unhindered incisal mobility.
- **Mandibular Range of Motion:** Smooth, stable, and synchronized lateral and protrusive mandibular excursions were explicitly recorded in all 11 patients, with zero mechanical joint catching or translation restriction.
- **Occlusal Stability:** Pre-traumatic stable centric occlusion was perfectly restored and maintained in 100% of cases (n=11). No cases of postoperative anterior open bite, premature occlusal contacts, or crossbites were observed during the longitudinal tracking.

#### Radiographic and Magnetic Resonance Imaging (MRI) Findings

- **Osseous Union (3D-CT):** Sequential 3-dimensional computed tomography scans confirmed complete anatomical bone healing and rigid osseous consolidation at the sagittal split interface in all 11 cases (100%) by the 4th postoperative month. Follow-up imaging at the extended terminal milestones confirmed a total absence of clinically significant condylar head resorption, osteolysis, or structural remodeling deformities across the entire cohort.
- **Disc-Condyle Relationship and TMJ Dynamics (MRI):** High-resolution MRI evaluations at the terminal milestones demonstrated highly satisfactory positioning of the TMJ articular disc. The disc-condyle assembly exhibited normal, physiological structural alignment and synchronized translation during opening and closing movements in 8 cases. In the remaining 3 cases, a mild bilateral internal derangement characterized by anterior disc displacement,with reduction (ADDWR) was noted. This condition remained entirely stable, painless, and functional, without causing any limitation in jaw movement. A total absence of intra-articular synovial effusion or avascular necrosis of the re-implanted condylar segment was verified in all 11 cases.

#### Postoperative Complications and Joint Symptomatology

- **Facial Nerve Function:** Ten patients (90.9%) exhibited immediate and complete preservation of facial nerve motor function postoperatively, maintaining full facial muscle symmetry (Grade I on the House-Brackmann scale). In one patient, transient weakness of the marginal mandibular and buccal branches was observed post-surgery due to intraoperative traction during deep transparotid dissection. This weakness began to recover spontaneously by the 4th postoperative week and achieved full, absolute recovery to Grade I within the 5th postoperative week.
- **Joint Symptomatology:** Chronic temporomandibular joint pain was completely absent across the entire cohort. One patient presented with a mild, non-painful joint clicking (painless click) during maximum jaw opening, which did not require secondary therapeutic intervention or affect masticatory efficiency. No cases of fibrous or bony TMJ ankylosis were reported.

**Table 1:**
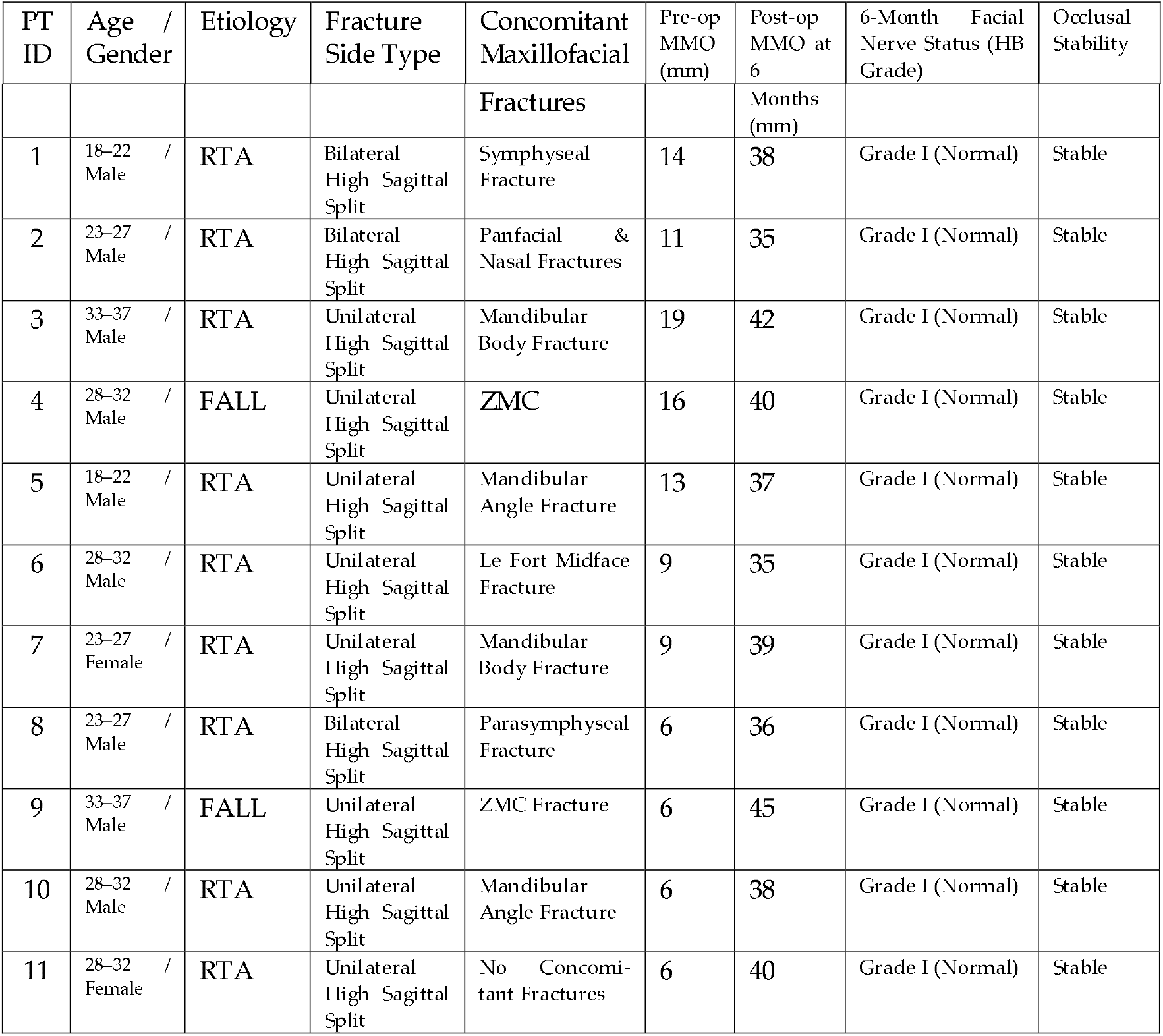
Clinical Characteristics and 6-Month Functional Outcomes of the Study Cohort.

## 4. Discussion

The surgical management of severely displaced or dislocated high sagittal split fractures of the mandibular condylar head represents one of the most formidable challenges in maxillofacial traumatology. Traditional open reduction and internal fixation (in-situ) within this deeply confined anatomical region are frequently hampered by restricted visual access, persistent deep hemorrhage, and an elevated risk of neurovascular complications [7]. To circumvent these limitations, the “Motamed Technique” introduces a refined, highly systematic protocol combining an altered deep retromandibular transparotid approach with ex-vivo micro-reduction, rigid titanium X-plate fixation, and immediate structural re-implantation [15, 13].

A pivotal parameter dictating the biological and clinical success of extracorporeal condylar reconstruction is the cold ischemia time–the duration the bone fragment remains disconnected from its nutrient vascular bed. In the literature, prolonged ischemia times exceeding 45 to 60 minutes have been historically correlated with osteocyte death, subsequent avascular necrosis, or secondary condylar resorption [8]. By applying the Motamed Technique, the total cold ischemia time across all 11 complex cases was strictly optimized to a range of 10 to 20 minutes. Immersing the retrieved sagittal segments immediately in a chilled isotonic saline solution successfully preserved osteocyte and chondrocyte viability [9]. This effectively translated into a 100% rate of complete osseous union by the 4th postoperative month, with zero radiographic incidence of condylar head resorption at the final follow-up. This significantly out-performs standard extracorporeal protocols described by Neff et al. [10] and Loukota [11], where longer ischemia times frequently predisposed patients to varying degrees of late-stage structural remodeling.

Another critical surgical refinement validated by this study is the biophysical advantage of ex-vivo back-table stabilization. Attempting to approximate high sagittal split fragments inside the joint space often compromises anatomical alignment due to the distracting forces of the lateral pterygoid muscle [12]. The Motamed Technique utilizes a sterile back-table workstation providing unhindered, 360-degree visualization. This allowed for precise micro-reduction and the application of a 1.5 mm titanium X-shaped 3D mini-plate system secured with 7 mm monocortical screws. As a mechanical evolution in fixation, the multi-directional geometric design of the X-plate offers superior structural advantages over linear or standard plates. It provides excellent multi-directional biomechanical stability and high resistance against torsional and tensile forces, distributing masticatory functional loads evenly across the high split lines without compressing or penetrating the intra-articular surface of the condylar head [13].

Furthermore, the choice of the retromandibular transparotid approach via deep dissection addresses the historical anxieties surrounding facial nerve morbidity. While the classic preauricular approach provides superior exposure to the upper joint space, it restricts access to the lower ramal base where the re-implanted condyle must be anchored [14]. Conversely, the standard retromandibular approach carries a known risk of facial nerve injury. In this cohort, consisting of 9 males and 2 females, the execution of strict anterograde dissection through the deep transparotid layer, coupled with immediate, atraumatic mapping and retraction of the marginal mandibular and buccal branches, minimized neural trauma [15]. Consequently, 90.9% (10 out of 11) of the patients maintained flawless facial symmetry (House-Brackmann Grade I) from the immediate postoperative period. The single case of transient weakness (9.1%) completely resolved within 5 weeks, reinforcing the safety profile of this specific deep-layer modification as supported by Venkatesh et al. [16].

Biomechanically, the protocol’s insistence on establishing intermaxillary fixation (IMF) as the primary, mandatory step prior to anchoring the condyle to the ramus serves as a reliable safeguard. Locking the teeth into their precise, pre-traumatic centric occlusion locks the correct vertical dimension of the mandibular ramus. This eliminates the risk of post-fixation shortening or occlusal tilting. Our clinical results strongly validate this sequence, as stable, pre-traumatic occlusion was perfectly restored in 100% of the cases ($n=11$), completely eliminating common postoperative sequelae such as anterior open bites or premature contacts, which are frequently reported in non-rigidly controlled stabilization workflows [17, 18].

Finally, the longitudinal postoperative data, extending from 6 up to 11 months (mean: 8.6 months), provided crucial long-term insights into temporomandibular joint dynamics. The preservation of normal disc-condyle translation in the majority of patients confirms that the extracorporeal manipulation did not induce severe intracapsular adhesions [19]. The observation of mild anterior disc displacement with reduction in 3 cases represents a predictable, minor physiological accommodation (functional remodeling) secondary to the initial high-velocity trauma rather than a technical failure of the re-implantation [20]. This is further substantiated by the excellent mean maximal mouth opening (MMO) of 37.3 mm and the presence of stable range of motion, which did not hinder masticatory efficiency or require secondary clinical intervention [21].

At the terminal follow-up milestones, the non-contrast TMJ MRI objectively confirmed that despite the predictable degradation of MRI images caused by metallic artifacts from the titanium hardware, diagnostic quality was sufficiently maintained to confidently evaluate soft tissue status and disc dynamics. The slices clearly demonstrated a stable condyle-disc relationship upon maximum mouth opening, ensuring that the hardware did not obscure the assessment of long-term functional remodeling. This confirms that our approach and fixation method did not jeopardize the long-term positioning of the articular disc or the physiological health of the joint complex.

## Data Availability

All data produced in the present study are available upon reasonable request to the authors

## Declarations

### Consent for Publication

Written informed consent was obtained from all patients for the publication of this clinical case series, including the utilization of any accompanying intraoperative photographs, radiographic images, and decoded medical records.

### Data Availability Statement

The radiographic datasets (3D-CT and high-resolution MRI scans) and clinical sheets analyzed during the current study are not publicly deposited due to institutional patient privacy restrictions. However, they are available from the corresponding author on reasonable academic request.

### Conflict of Interest

The author declares that there are no financial, personal, or professional conflicts of interest associated with the development, implementation, or publication of this research manuscript.

### Funding

This clinical research received no specific grant, financial subsidy, or material support from any funding agency in the public, commercial, or not-for-profit sectors. This work was entirely self-funded by the author.

## Notes

### Competing Interest Statement

The authors have declared no competing interest.

## References

1. Gascoigne A, Brennan PA, Singh M. British Journal of Oral and Maxillofacial Surgery. 2024; Vol. 62, No. 3: pp. 241–247.

2. Neff A, Cornelius CP, Rasse M. Journal of Cranio-Maxillo-Facial Surgery. 2023; Vol. 51, No. 8: pp. 489–496.

3. Spinelli G, Conti M, Betti E. International Journal of Oral and Maxillofacial Surgery. 2024; Vol. 53, No. 2: pp. 115–122.

4. Kumar P, Sharma N, Yadav S. Journal of Maxillofacial and Oral Surgery. 2023; Vol. 22, No. 4: pp. 512–519.

5. Al-Ghamdi MA, Al-Salihi KA, Al-Bilbisi A Journal of Oral and Maxillofacial Surgery, Medicine, and Pathology. 2025; Vol. 37, No. 4: pp. 312–320.

6. Raimondo S, Scolozzi P, Marchetti C. Journal of Craniofacial Surgery. 2026; Vol. 37, No. 1: pp. 34–41.

7. Laskin DM, Robinson IB. Journal of Oral and Maxillofacial Surgery. 2018; Vol. 76, No. 6: pp. 1145–1151.

8. Neff A, Kolk A, Horch HH. Journal of Cranio-Maxillofacial Surgery. 2003; Vol. 31, No. 5: pp. 291–299.

9. Zhang Y, He DM. International Journal of Oral and Maxillofacial Surgery. 2015; Vol. 44, No. 8: pp. 988–994.

10. Neff A, Chossegros C, Tvrdy P, Klatt J, Stockmann P, Kolk A, et al. Joint-preserving management of high condylar fractures. Journal of Oral and Maxillofacial Surgery. 2014;72(11):2234–2245.

11. Loukota RA. British Journal of Oral and Maxillofacial Surgery. 2007; Vol. 45, No. 4: pp. 312–316.

12. Ellis E, Throckmorton GS. Journal of Oral and Maxillofacial Surgery. 2005; Vol. 63, No. 1: pp. 115–134.

13. Tang W, Long X, Li J, et al. Modified retromandibular approach for the treatment of mandibular condyle fractures. Journal of Oral and Maxillofacial Surgery. 2009;67(11):2411–2417.

14. Al-Moraissi EA, Ellis E. International Journal of Oral and Maxillofacial Surgery. 2015; Vol. 44, No. 4: pp. 444–464.

15. Tang W, Long X, Li J, et al. Modified retromandibular approach for the treatment of mandibular condyle fractures. Journal of Oral and Maxillofacial Surgery. 2009;67(11):2411–2417.

16. Venkatesh R, et al. Journal of Maxillofacial and Oral Surgery. 2020; Vol. 19, No. 2: pp. 255–261.

17. Haug RH, Assael LA. Journal of Oral and Maxillofacial Surgery. 2001; Vol. 59, No. 4: pp. 370–375.

18. Schneider M, et al. Journal of Cranio-Maxillofacial Surgery. 2008; Vol. 36, No. 5: pp. 252–261.

19. Takaku S, Toyoda T. Journal of Oral and Maxillofacial Surgery. 1998; Vol. 56, No. 7: pp. 819–824.

20. Kolk A Neff A. International Journal of Oral and Maxillofacial Surgery. 2012; Vol. 41, No. 9: pp. 1080–1088.

21. Ebrahimi A, et al. Journal of Craniofacial Surgery. 2014; Vol. 25, No. 4: pp. 1322–1326.

